# Cross-Ancestry Remapping of the Chromosome 1q31 Th2 pathway-associated interval Refines an Asthma Association Signal in Patients with Steroid-Dependent Disease

**DOI:** 10.64898/2026.05.06.26352550

**Authors:** Hui-Qi Qu, Haijun Qiu, Frank D Mentch, Christopher J Cardinale, Hakon Hakonarson

## Abstract

**Background:** The chromosome 1q31 Th2 pathway-associated interval has been linked to asthma, but its phenotype specificity and cross-ancestry architecture remain unclear.

**Methods:** We analyzed African (AFR) and European (EU) ancestry datasets, including 9,965 asthma cases and 37,391 controls of AFR, and 6,074 cases and 116,255 controls of EU ancestry. Imputed dosage-based association analyses were performed for asthma, steroid-dependent asthma (SDA), and non-steroid-dependent asthma, followed by QC-filtered SDA remapping, leave-one-batch-out analysis, cross-ancestry comparison, and functional enrichment.

**Results:** Strong regional association was observed only for SDA. After quality-control (QC) filtering, the SDA signal remained significant in both ancestries, with 2,280 genome-wide significant variants in AFR and 859 in EU. Cross-ancestry comparison identified 3,129 significant variants: 10 shared, 2,270 AFR-specific, and 849 EU-specific. Shared variants showed concordant effects, whereas 237 variants showed nominal heterogeneity. AFR-specific signals included *PTPRC* variants with larger effects in AFR. Functional enrichment suggested different biological emphases within the same interval: immune and contractile airway-wall biology in AFR, and additional neuroaxonal components in EU.

**Conclusions:** The 1q31 interval is strongly associated with SDA in both AFR and EU populations, and its fine-scale architecture differs by ancestry. These findings highlight population-specific effects within a shared SDA susceptibility interval, with potential implications for population-informed precision medicine in steroid responsiveness and asthma management.

## Introduction

Genetic heterogeneity in asthma can obscure locus-specific effects in broad case-control analyses. Although genome-wide association studies (GWAS) have identified many reproducible asthma loci, biological interpretation remains difficult because most associated variants are noncoding and lie within regional haplotypes containing multiple correlated markers and candidate genes^1, 2^. Phenotype refinement and cross-ancestry comparison offer complementary strategies for improving interpretation of established loci. The chromosome 1q31 T helper 2 (Th2) pathway-associated interval is an example. In our earlier work, variation in this region was associated with steroid-dependent asthma (SDA) and showed differences between African (AFR)- and European (EU)-ancestry groups^3^. The interval includes *DENND1B*, and subsequent functional studies showed that *DENND1B* regulates T-cell receptor signaling in Th2 cells, linking the locus to allergic airway inflammation^4^. However, the broader 1q31 interval contains additional genes with plausible immunologic relevance, suggesting that the association may reflect a regional regulatory domain rather than the effect of a single gene. Because SDA is commonly associated with type 2 inflammatory features^5^, this subphenotype may provide a more specific context for resolving the association signal.

Cross-ancestry analysis can further clarify whether an established locus has the same genetic architecture across populations or contains population-specific effects. Differences in allele frequency, haplotype background, regulatory architecture, environmental exposures, and gene-environment interactions may cause the same regional locus to show different variant-level association patterns across ancestry groups^6, 7^. Comparing AFR and EU datasets can distinguish a broadly shared association from ancestry-specific signals within the same locus. We revisited the chromosome 1q31 interval in African- and European-ancestry datasets using dense imputation, dosage-based association testing, and phenotype stratification by requirement for chronic high-dose inhalded or oral steroid use. We compared association patterns for asthma, SDA, and non-steroid-dependent asthma (NSDA), followed by quality-controlled regional remapping of the SDA phenotype and cross-ancestry comparison. Our aim was to determine whether 1q31 shows a shared SDA association across populations and how variant-level patterns differ between AFR and EU.

## Methods

### Study design

This study was conducted separately in African (AFR) and European (EU) ancestry subsets. Genotype data were analyzed as imputed dosages rather than best-guess genotype calls. The AFR dataset comprised 37 genotyping array or imputation batches, including 9,965 asthma cases and 37,391 controls. The EU dataset comprised 38 genotyping array or imputation batches, including 6,074 asthma cases and 116,255 controls. Genome-wide association analyses were first performed for all asthma cases versus controls, followed by stratified analyses of SDA (AFR, 116 cases; EU, 186 cases) and NSDA (AFR, 9,849 cases; EU, 5,888 cases). The SDA cohort included patients with persistent asthma of moderate to high severity who required high doses of daily inhaled glucocorticoid therapy or oral steroids to control their asthma symptoms. The SDA and NSDA phenotypes were established through comprehensive electronic health record (EHR) data review with a minimum of 3 years of longitudinal data, including multiple patient visits and tracking of inhaled or oral steroid use. Patients meeting criteria for chronic high-dose inhaled glucocorticoid therapy or oral steroid requirement were classified as SDA, whereas asthma cases without this requirement were classified as NSDA. Sample details are provided in Supplementary Tables 1 and 2.

### Genotyping, quality control, and imputation

Samples were genotyped on multiple Illumina arrays, including Global Screening Array, HumanHap, Omni25, and OmniExpress/OmniExpressExome-based merged subsets (Illumina, CA, USA). Genotype data were processed in batch-specific datasets corresponding to genotyping array or imputation subsets. Variant alleles were harmonized to the reference panel before imputation. Genome-wide imputation was performed using the TOPMed Imputation Server (version R2, GRCh38)^8^.

### Genetic association analysis

Genetic association analyses were performed separately within the EU and AFR ancestry subsets using PLINK v2.0 on imputed dosage data. Within each ancestry group, principal component analysis (PCA) was conducted using ancestry-specific quality-controlled genotype data. The first six principal components were included as covariates in downstream association analyses.

Analyses were performed for three binary phenotypes: asthma, SDA, and NSDA. For each ancestry group and genotyping array or imputation batch, association testing was performed using PLINK 2 --glm, with adjustment for age, sex, and the first six ancestry-specific principal components. Covariates were variance-standardized before model fitting to improve numerical stability, and association results were generated separately for each batch and ancestry group. Because the outcomes were binary, association testing used logistic regression with Firth fallback where needed.

### Imputation-batch-level meta-analysis

Imputation-batch-specific summary statistics were filtered to retain additive genetic effect results and were then combined across imputation batches within ancestry using inverse-variance weighted fixed-effects meta-analysis. Each imputation batch was treated as an independent analysis unit, including multiple batches derived from the same genotyping array, because imputed dosages incorporate batch-specific uncertainty from imputation quality metrics (e.g., INFO or R²) ^9^. Meta-analyses were performed separately for each phenotype within the EU and AFR datasets.

### Remapping of the Th2 pathway locus

To further interrogate the association signal identified in the genome-wide analyses, we performed a region-focused follow-up analysis of the Th2 pathway-associated interval on chromosome 1 (chr1:197,356,770-205,766,770; GRCh38). This analysis used ancestry-stratified batch-level association results extracted from the genome-wide output and was performed separately in AFR and EU participants for the SDA phenotype. For the regional analysis, additional batch-level quality-control filters were applied, requiring at least 2 cases per batch and representation of both sexes among cases; batches failing either criterion were excluded. For each retained batch, regional association files were harmonized to a common allele definition by parsing the reference and alternate alleles from the variant identifier and aligning all study-specific effects to the alternate allele, reversing effect estimates when necessary.

Within each ancestry, variants in the target interval were recombined across retained batches using fixed-effect inverse-variance weighted meta-analysis. For each variant, we calculated the pooled effect estimate, standard error, z statistic, and meta-analysis p value using the one-degree-of-freedom chi-square survival function. Between-batch heterogeneity was summarized using Cochran’s Q, its p value, and I^2^. To assess robustness to batch composition, we performed a leave-one-batch-out sensitivity analysis in which the regional meta-analysis was repeated after excluding each retained batch in turn.

Regional SDA association results from AFR and EU analyses were compared for variants reaching genome-wide significance in at least one ancestry group. Variants significant in both ancestry groups were classified as shared, whereas variants significant in only one ancestry group were classified as AFR-specific or EU-specific. Cross-ancestry fixed-effects meta-analysis was then performed from the ancestry-specific effect estimates and standard errors, and between-ancestry heterogeneity was assessed with Cochran’s Q and I^2^. Variants with heterogeneity p < 0.05 were considered to show nominal evidence of cross-ancestry effect-size heterogeneity.

### Functional annotation

Genome-wide significant variants from the cross-ancestry comparison were grouped as shared, AFR-specific, or EU-specific. Genes overlapping these signals were identified by positional mapping to Ensembl gene annotations, and enrichment analysis was performed separately for each gene set using g:Profiler ^10^ with false discovery rate correction. Enriched terms were compared across ancestry-specific and shared signal classes to identify biological features common to both populations and those that were population-specific.

## Results

### Phenotype-specific association

We performed regional remapping in ancestry-specific meta-analyses of overall asthma, SDA, and NSDA across the previously implicated chromosome 1q31 interval spanning 8.4 Mb. This interval spans the three previously identified markers rs2786098, rs2014202, and rs7522056 (GRCh38 positions chr1:197,356,778, chr1:205,766,484, and chr1:205,766,763, respectively). The interval includes *DENND1B* and several additional genes with plausible immune or inflammatory relevance, including *NEK7*, *PTPN7, ADORA1, CHI3L1, CHIT1*, and the *MIR181A/B* locus.

Across the overall asthma meta-analyses, only modest evidence of association was observed in both ancestry groups (Table 1; Figure 1). In AFR, the lead signal for overall asthma was chr1:204034569:T:C (P = 1.54E-5; β = 0.096), whereas in EU the lead signal was chr1:201189027:G:A (P = 8.95E-6; β = 0.388). No variants reached genome-wide significance in either ancestry group for overall asthma. Similarly, NSDA showed only suggestive evidence of association in both ancestry groups (Table 1; Figure 1). The lead variant in AFR was chr1:198404112:G:A (P = 6.20E-5; β = 0.086), whereas the lead variant in EU was chr1:202976495:C:T (P = 5.05E-6; β = 0.272). As in the overall asthma analyses, no variants exceeded genome-wide significance for NSDA in either ancestry group.

**Figure 1.**
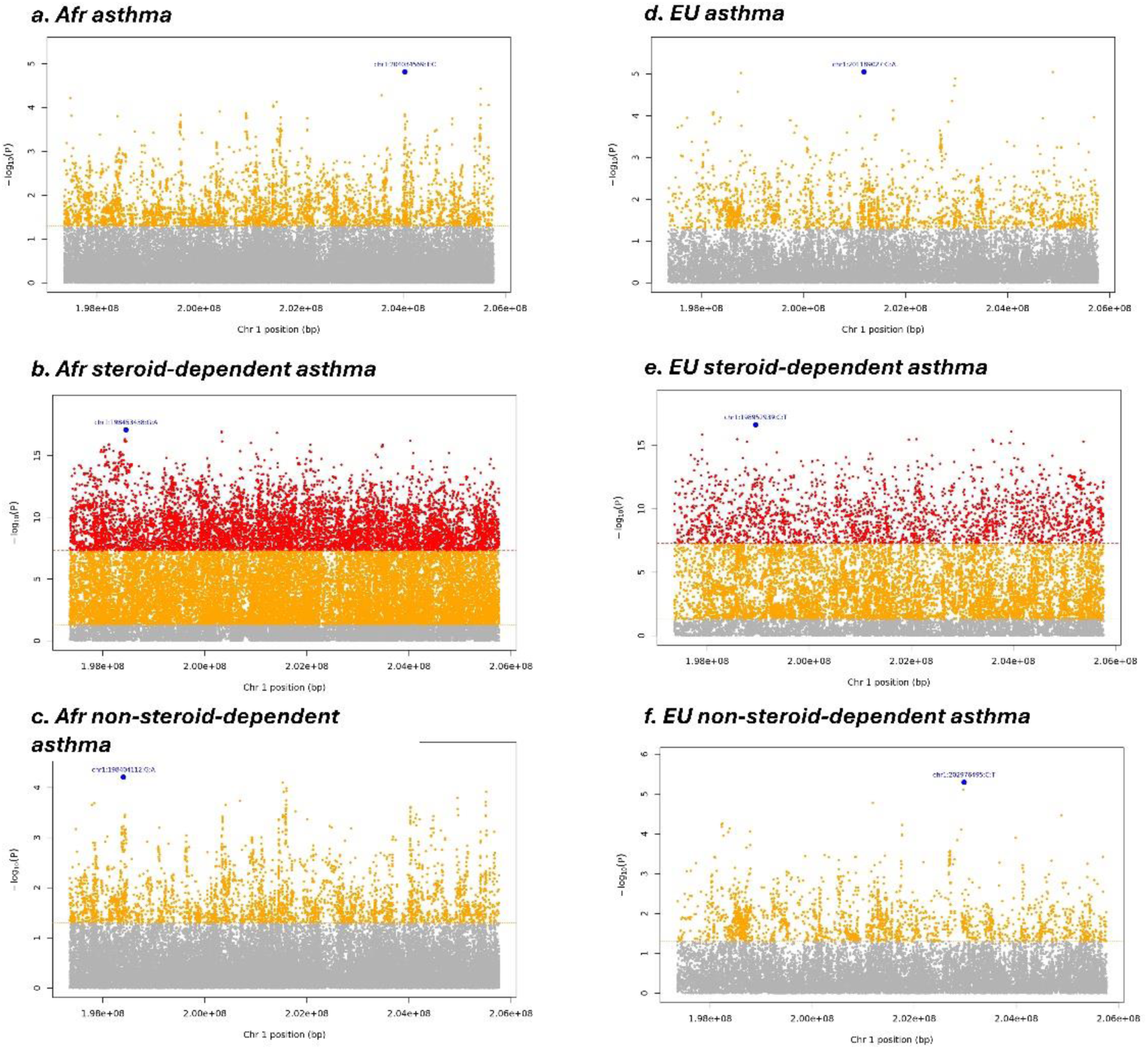
Regional Manhattan plots for the Th2 pathway–associated region on chromosome 1 (197,356,770–205,766,770 bp) from each meta-analysis. Each point represents one variant, plotted by genomic position on the x-axis and association significance as −log10(P) on the y-axis. Variants reaching genome-wide significance (P < 5E-8) are shown in red. Variants with nominal significance (5E-8 ≤ P < 0.05) are shown in orange. Variants with P ≥ 0.05 are shown in gray. The red dashed horizontal line marks the genome-wide significance threshold (P = 5E-8), and the orange dashed horizontal line marks the nominal significance threshold (P = 0.05). Panels show (a) EU asthma, (b) EU SDA, (c) EU NSDA, (d) AFR asthma, (e) AFR SDA, and (f) AFR NSDA.

**Table 1.**
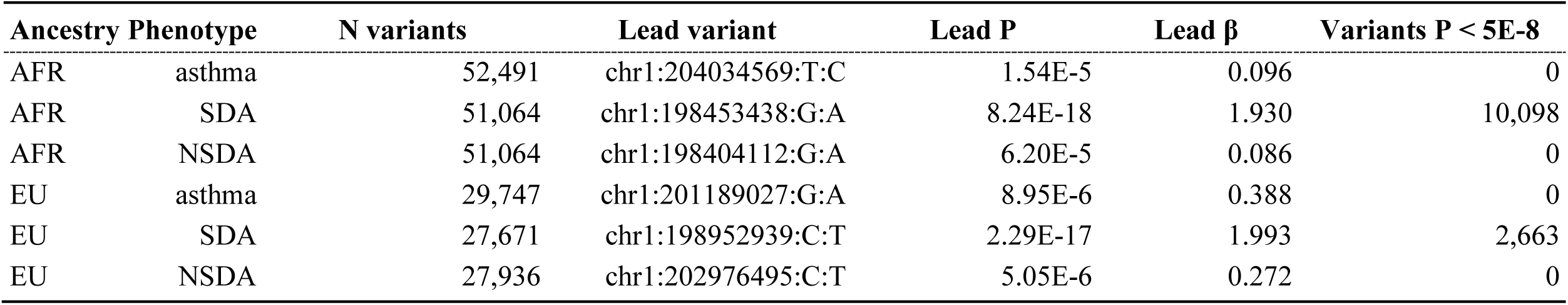
Regional associations across the Th2 pathway–associated region.

In contrast, strong evidence of association across the region was observed for SDA in both ancestry groups (Table 1; Figure 1). In AFR, the lead variant was chr1:198453438:G:A (P = 8.24E-18; β = 1.930), with 10,098 variants exceeding genome-wide significance within the interval. In EU, the lead variant was chr1:198952939:C:T (P = 2.29E-17; β = 1.993), with 2,663 variants exceeding genome-wide significance (Table 1). Regional Manhattan plots showed dense clusters of highly significant variants across multiple segments of the interval in SDA, whereas the corresponding overall asthma and NSDA analyses showed only scattered suggestive signals (Figure 1).

### QC-filtered regional follow-up analysis of SDA

Given that the strongest regional signal was observed in SDA, we next performed a QC-filtered follow-up analysis focused on this phenotype. After imputation-batch-level quality control requiring at least 2 SDA cases per batch and representation of both sexes among cases, 23 AFR batches and 27 EU batches were retained for the SDA regional analysis.

Using these QC-filtered data, the regional association remained strongly associated with SDA in both ancestries (Table 2; Figure 2). In AFR, the lead variant remained chr1:198453438:G:A (P = 2.28E-14; β = 1.878), with 2,280 variants reaching P < 5E-8. In EU, 27,553 variants were meta-analyzed, and the lead variant was chr1:203600088:T:TA (P = 2.41E-14; β = 1.976), with 859 variants reaching P < 5E-8 (Table 2). Thus, although the number of genome-wide significant variants was reduced after application of the additional imputation-batch-level filters, the regional signal remained highly significant in both ancestry groups.

**Figure 2.**
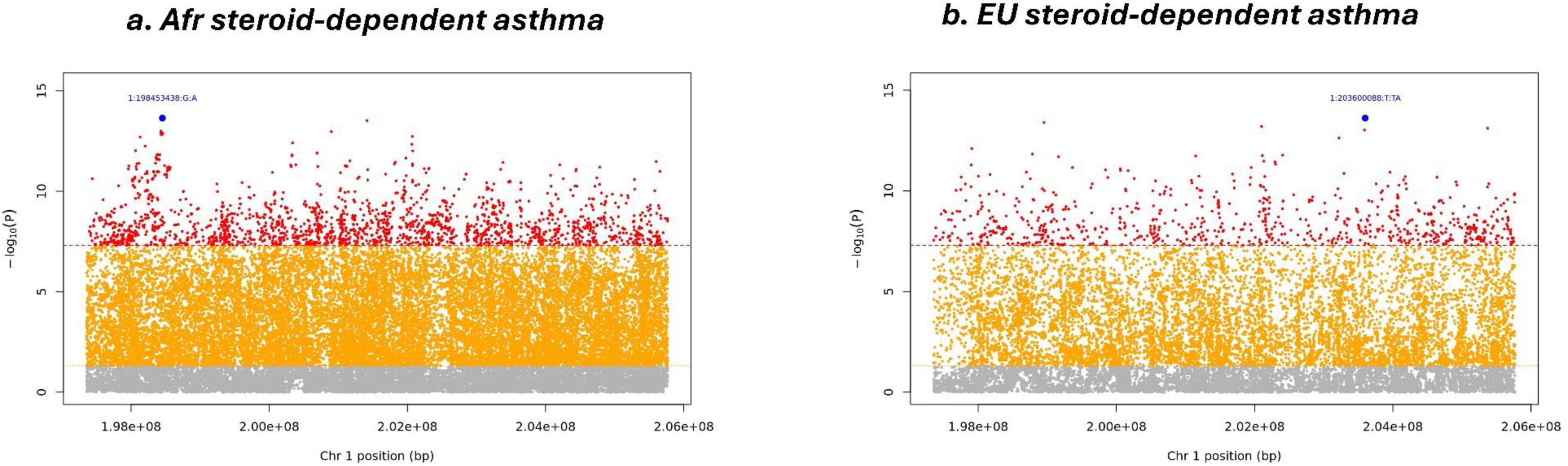
QC-filtered regional Manhattan plots for SDA in the Th2 pathway–associated region. Regional meta-analysis results for SDA after chip-level quality control requiring at least 2 cases per chip and representation of both sexes among cases. Panels show (a) EU and (b) AFR ancestry groups. Each point represents one variant, plotted by genomic position on the x-axis and association significance as −log10(P) on the y-axis. Variants reaching genome-wide significance (P < 5E-8) are shown in red, variants with nominal significance (5E-8 ≤ P < 0.05) in orange, and variants with P ≥ 0.05 in gray. The red dashed horizontal line marks the genome-wide significance threshold, and the orange dashed horizontal line marks the nominal significance threshold.

**Table 2.** QC-filtered regional associations across the Th2 pathway–associated region for SDA.

| Ancestry | N chips included | N variants | Lead variant | Lead P | Lead $\beta$ | Variants P < 5E-8 |
| --- | --- | --- | --- | --- | --- | --- |
| AFR | 23 | 50,587 | chr1:198453438:G:A | 2.28E-14 | 1.878 | 2,280 |
| EU | 27 | 27,553 | chr1:203600088:T:TA | 2.41E-14 | 1.976 | 859 |

Heterogeneity at the lead variant was minimal in both ancestries. In AFR, the lead signal was supported by 23 contributing imputation batches, with I^2^ = 0.0 and Cochran’s Q P = 0.977. In EU, the lead signal was supported by 25 contributing imputation batches, with I^2^ = 0.0 and Q P = 0.917. These findings indicate consistency of the lead association across retained batches within each ancestry group.

### Leave-one-batch-out sensitivity analysis

To evaluate whether the regional association was disproportionately driven by any single imputation batch, we performed leave-one-batch-out sensitivity analyses across all retained batches (Table 3). In AFR, 23 leave-one-batch-out runs were completed. Across these runs, the lead P value ranged from 1.59E-14 to 5.21E-13, the number of genome-wide significant regional variants ranged from 1,475 to 2,086. The main-analysis lead variant, chr1:198453438:G:A, remained the lead in 12 of 23 runs. In EU, 27 leave-one-batch-out runs were completed. The lead P value ranged from 8.41E-15 to 2.34E-13, the number of genome-wide significant regional variants ranged from 519 to 834. The main-analysis lead variant, chr1:203600088:T:TA, remained the lead in 18 of 27 runs.

**Table 3.** Leave-one-chip-out sensitivity analysis of the QC-filtered regional meta-analysis of the Th2 pathway–associated region.

| Ancestry | N leave-one-chip-out runs | Main lead variant | Lead P range | Runs retaining main lead | Variants P < 5E-8 range |
| --- | --- | --- | --- | --- | --- |
| AFR | 23 | chr1:198453438:G:A | 1.59E-14 to 5.21E-13 | 12/23 | 1,475 to 2,086 |
| EU | 27 | chr1:203600088:T:TA | 8.41E-15 to 2.34E-13 | 18/27 | 519 to 834 |

Inspection of the leave-one-batch-out runs showed that, although the exact lead variant could shift, the overall regional signal persisted after exclusion of any single batch in both ancestries. Thus, the SDA association across the chromosome 1q31 Th2 pathway-associated interval was not dependent on any single contributing imputation batch, although fine-scale localization of the lead signal varied modestly with batch composition.

### QC-filtered SDA regional associations

In the AFR ancestry meta-analysis, we identified four genome-wide significant variants at chromosome 1q31 within the *PTPRC* locus (1:198684045:A:G, 1:198707429:GC:G, 1:198754608:G:A, and 1:198760968:ATG:A; all P < 5E-8) that correspond to previously reported GWAS signals, including findings from UK Biobank-based studies (Table 4). Effect sizes were consistently large in African ancestry samples (β range: 1.39-1.62), whereas the corresponding effects in European ancestry were smaller (β range: 0.33-0.57) and did not reach genome-wide significance. Cross-ancestry meta-analysis remained significant for all four variants, but heterogeneity tests indicated substantial differences between ancestries (all heterogeneity P ≤ 0.0011).

**Table 4.** African ancestry genome-wide significant variants at the *PTPRC* locus and comparison with European ancestry and prior GWAS evidence.

| Variant* | Consequence | Gene | Effect allele | FREQ_AFR | AFR meta-analysis | EU meta-analysis | Cross-ancestry result | Prior GWAS evidence |
| --- | --- | --- | --- | --- | --- | --- | --- | --- |
| chr1:198684045:A:G | intron | *PTPRC* | G | 0.0216 | $\beta = 1.39$, $P = 4.41\text{E-}9$ | $\beta = 0.33$, $P = 0.065$ | $P = 6.14\text{E-}7$; heterogeneity $P = 3.14\text{E-}4$ | Reported in prior GWASs including UK Biobank<sup>14-17</sup> |
| chr1:198707429:GC:G | intron | *PTPRC* | G | 0.0137 | $\beta = 1.62$, $P = 2.69\text{E-}8$ | $\beta = 0.48$, $P = 0.010$ | $P = 2.29\text{E-}7$; heterogeneity $P = 0.0010$ | Previously reported; lead SNP in prior GWAS<sup>14</sup> |
| chr1:198754608:G:A | intron | *PTPRC* | A | 0.0171 | $\beta = 1.54$, $P = 1.04\text{E-}10$ | $\beta = 0.57$, $P = 0.0012$ | $P = 1.21\text{E-}10$; heterogeneity $P = 0.0011$ | Reported in prior GWAS<sup>14</sup> |
| chr1:198760968:ATG:A | downstream_gene | *PTPRC* | A | 0.0171 | $\beta = 1.55$, $P = 9.93\text{E-}11$ | $\beta = 0.57$, $P = 0.0012$ | $P = 1.15\text{E-}10$; heterogeneity $P = 0.0010$ | Reported in prior GWAS<sup>14</sup> |

\*These variants showed strong LD in both AFR and EU. chr1:198754608:G:A and chr1:198760968:ATG:A are in near-complete LD in both populations (mean $r^2 = 0.997$ in AFR; 0.998 in EU). The remaining pairwise correlations ranged from mean $r^2 = 0.585$ to 0.697 in AFR and 0.647 to 0.931 in EU.

We compared the QC-filtered regional association results for SDA between the AFR and EU ancestry analyses. Ten variants reached genome-wide significance in both ancestry groups (Table 5). These shared variants were located at intergenic positions and within KIF14 and GPR37L1, and all showed concordant positive effect directions in AFR and EU. Effect sizes were also similar across ancestries for most of these variants, with heterogeneity P values ranging from 0.2122 to 0.8309 and I^2^ = 0 for nine of the ten variants. These findings indicate that, despite the predominance of ancestry-specific significant variants across the interval, a small core set of variants shows consistent association with SDA across both ancestry groups.

**Table 5.**
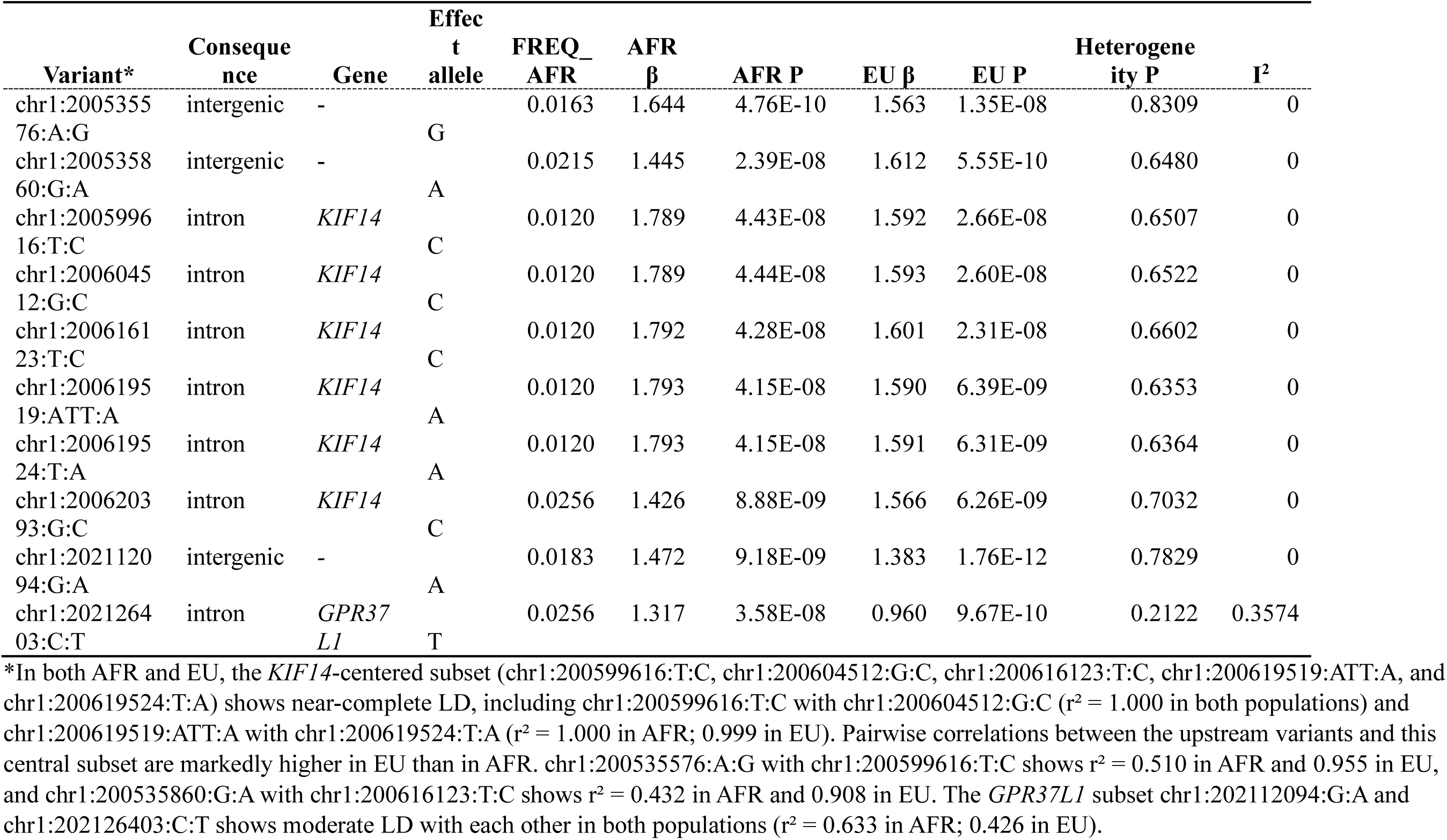
Genome-wide significant variants in both African and European.

Of the genome-wide significant variants, 2,270 were significant only in AFR and 849 only in EU. In total, 3,129 unique variants reached genome-wide significance in at least one ancestry group across the interval. Cross-ancestry heterogeneity testing identified 237 variants with nominal evidence of between-ancestry effect-size differences (Q P < 0.05), including 147 AFR-specific variants and 90 EU-specific variants. In particular, six variants showed opposite effect directions between AFR and EU after allele harmonization (Table 6). These variants mapped to *LHX9, SYT2, ADORA1*, and *ELK4*, with two additional intergenic signals. Of the six, three were AFR-only genome-wide significant variants with positive effects in AFR and negative effects in EU, two were EU-only genome-wide significant variants with negative effects in EU and positive effects in AFR, and one additional AFR-only variant at ADORA1 showed an opposite direction but little evidence of cross-ancestry heterogeneity. For five of the six variants, heterogeneity was substantial (heterogeneity P ranging from 6.42 x 10⁻¹³ to 9.93 x 10⁻⁸; I^2^ = 0.964-0.981), supporting genuine ancestry-dependent differences in local signal configuration. These findings indicate that a subset of regional associations is not only ancestry-specific in significance but also opposite in effect direction across populations.

**Table 6.**
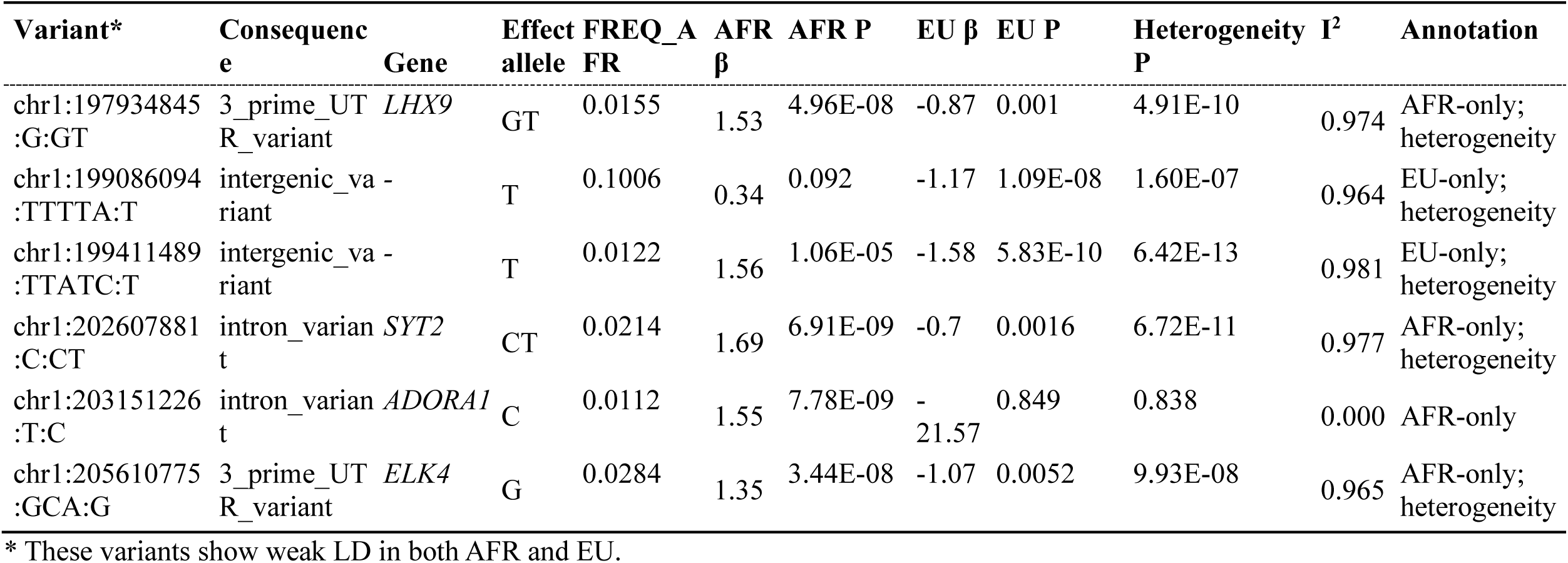
Variants with opposite AFR and EU effect directions after allele harmonization.

To assess whether ancestry-specific signals mapped to similar or distinct biological annotations, we performed functional overrepresentation analysis of genes mapped from AFR-specific, EU-specific, and shared significant variants. The AFR-specific and EU-specific sets comprised 110 and 92 genes, respectively, whereas the shared set comprised only 2 genes and was not examined further because of its small size. The AFR-specific and EU-specific gene sets showed overlapping enrichment profiles (Table 7). In both sets, the top enriched terms included myofibril, sarcomere, contractile muscle fiber, supramolecular polymer, and supramolecular fiber. The EU-specific set also included axon initial segment, main axon, and I band.

**Table 7.** Representative significant enrichment results from cross-ancestry comparison of QC-filtered SDA regional associations.

| Comparison class | Source | Term | P value | Genes |
| --- | --- | --- | --- | --- |
| AFR-specific gene set | GO:CC | myofibril | 2.03E-05 | *CACNA1S, IGFN1, TNNT2, TNNI1, CSRP1, LMOD1, PPP1R12B, MYBPH, ATP2B4* |
| AFR-specific gene set | GO:CC | sarcomere | 2.03E-05 | *CACNA1S, IGFN1, TNNT2, TNNI1, CSRP1, LMOD1, PPP1R12B, MYBPH, ATP2B4* |
| AFR-specific gene set | GO:CC | contractile muscle fiber | 2.03E-05 | *CACNA1S, IGFN1, TNNT2, TNNI1, CSRP1, LMOD1, PPP1R12B, MYBPH, ATP2B4* |
| AFR-specific gene set | GO:CC | supramolecular polymer | 1.51E-04 | *NEK7, KIF14, CAMSAP2, KIF21B, CACNA1S, IGFN1, PKP1, TNNT2, TNNI1, CSRP1, NAV1, LMOD1, PPP1R12B, MYBPH, ATP2B4* |
| AFR-specific gene set | GO:CC | supramolecular fiber | 1.51E-04 | *NEK7, KIF14, CAMSAP2, KIF21B, CACNA1S, IGFN1, PKP1, TNNT2, TNNI1, CSRP1, NAV1, LMOD1, PPP1R12B, MYBPH, ATP2B4* |
| EU-specific gene set | GO:CC | myofibril | 7.29E-05 | *CACNA1S, IGFN1, TNNI1, CSRP1, LMOD1, PPP1R12B, MYBPH, ATP2B4* |
| EU-specific gene set | GO:CC | sarcomere | 7.29E-05 | *CACNA1S, IGFN1, TNNI1, CSRP1, LMOD1, PPP1R12B, MYBPH, ATP2B4* |
| EU-specific gene set | GO:CC | contractile muscle fiber | 7.29E-05 | *CACNA1S, IGFN1, TNNI1, CSRP1, LMOD1, PPP1R12B, MYBPH, ATP2B4* |
| EU-specific gene set | GO:CC | supramolecular polymer | 1.17E-04 | *NEK7, KIF14, CAMSAP2, KIF21B, CACNA1S, IGFN1, PKP1, TNNI1, CSRP1, NAV1, LMOD1, PPP1R12B, MYBPH, ATP2B4* |
| EU-specific gene set | GO:CC | supramolecular fiber | 1.17E-04 | *NEK7, KIF14, CAMSAP2, KIF21B, CACNA1S, IGFN1, PKP1, TNNI1, CSRP1, NAV1, LMOD1, PPP1R12B, MYBPH, ATP2B4* |
| EU-specific gene set | GO:CC | axon initial segment | 2.04E-03 | *NAV1, NFASC, CNTN2* |
| EU-specific gene set | GO:CC | main axon | 2.19E-03 | *NAV1, ADORA1, NFASC, CNTN2* |
| EU-specific gene set | GO:CC | I band | 2.94E-03 | *CACNA1S, IGFN1, CSRP1, PPP1R12B, ATP2B4* |
| Cross-ancestry heterogeneous set | GO:CC | myofibril | 3.73E-04 | CACNA1S, IGFN1, TNNT2, CSRP1, PPP1R12B, ATP2B4 |
| Cross-ancestry heterogeneous set | GO:CC | contractile muscle fiber | 3.73E-04 | CACNA1S, IGFN1, TNNT2, CSRP1, PPP1R12B, ATP2B4 |
| Cross-ancestry heterogeneous set | GO:CC | sarcomere | 3.73E-04 | CACNA1S, IGFN1, TNNT2, CSRP1, PPP1R12B, ATP2B4 |
| Cross-ancestry heterogeneous set | GO:CC | I band | 3.73E-04 | CACNA1S, IGFN1, CSRP1, PPP1R12B, ATP2B4 |
| Cross-ancestry heterogeneous set | GO:CC | axon initial segment | 3.88E-04 | NAV1, NFASC, CNTN2 |
| Cross-ancestry heterogeneous set | GO:CC | supramolecular polymer | 3.88E-04 | NEK7, KIF14, CACNA1S, IGFN1, PKP1, TNNT2, CSRP1, NAV1, PPP1R12B, ATP2B4 |
| Cross-ancestry heterogeneous set | GO:CC | supramolecular fiber | 3.88E-04 | NEK7, KIF14, CACNA1S, IGFN1, PKP1, TNNT2, CSRP1, NAV1, PPP1R12B, ATP2B4 |

Because population-specific significance and cross-ancestry heterogeneity reflect different features of the signal, we analyzed heterogeneous variants separately. The set of variants showing nominal cross-ancestry heterogeneity and genome-wide significance in at least one ancestry mapped to 50 genes. Functional overrepresentation analysis of this heterogeneous set identified enrichment for myofibril, sarcomere, contractile muscle fiber, I band, axon initial segment, supramolecular polymer, and supramolecular fiber. These results place the heterogeneous set within a partially overlapping group of structural and axon-related cellular component terms.

## Discussion

### SDA refines the 1q31 association signal

The present study identifies SDA as the phenotype in which the chromosome 1q31 Th2 pathway-associated interval shows its strongest association, and demonstrates that this signal has extensive population-specific architecture. Although the interval was significantly associated with SDA in both AFR and EU ancestry groups, only 10 genome-wide significant variants were shared across ancestries, compared with 2,270 AFR-specific and 849 EU-specific variants. In addition, 237 variants showed nominal cross-ancestry heterogeneity, including variants with opposite effect directions. These findings indicate that the main distinction between AFR and EU is not simply the presence or absence of a 1q31 association, but the fine-scale configuration of association signals within the same SDA locus.

After additional batch-level quality control, the SDA association remained highly significant in both ancestry groups. Leave-one-batch-out analyses supported the same conclusion, and heterogeneity at the lead variant was minimal within each ancestry, indicating that the main association was not driven by inconsistent effects across contributing batches. These analyses show that the regional signal is robust, while its fine-scale localization differs substantially by ancestry. This degree of asymmetry is too large to dismiss as minor variation in lead-marker ranking.

Corticosteroid dependency in asthma is strongly linked to Th2-driven inflammation, including IL-4, IL-5, and IL-13 signaling, eosinophilia, and IgE-mediated responses ^1, 11^. The restriction of the 1q31 signal to SDA therefore places the locus within a well-defined immunological axis rather than within the heterogeneous umbrella of asthma diagnosis. For the shared *KIF14* and *GPR37L1* signals in AFR and EU, these genes may connect the SDA association to tissue responses downstream of Th2 inflammation. *KIF14* is a dual microtubule/F-actin binding protein involved in cytokinesis, providing a plausible link to airway remodeling in SDA^12^. *GPR37L1* encodes a G protein-coupled receptor with reported roles in neural and glial biology, which may relate to neuroimmune regulation of airway hyperresponsiveness^13^.

The *PTPRC* signal adds an important layer to this interpretation. Four genome-wide significant variants within *PTPRC* were identified in AFR, with large effect sizes and prior GWAS support ^14–17^, whereas the corresponding effects in EU were smaller, did not reach genome-wide significance, and showed cross-ancestry heterogeneity. *PTPRC* encodes CD45, a receptor-type protein tyrosine phosphatase that regulates antigen-receptor signaling thresholds in lymphocytes and thereby influences T-cell activation, Th2 polarization, and cytokine output ^18^. Gene-environment interaction is a plausible hypothesis that could contribute to the AFR-EU heterogeneity, as the effect of *PTPRC* variation on asthma risk may be amplified by exposures that repeatedly stimulate immune responses, including viral infections, allergen burden, air pollution, and socioeconomic stress. Therefore, the 1q31 interval contains multiple Th2-relevant regulatory components, including our previously reported *DENND1B*-mediated T-cell receptor signaling ^4^, as well as the prior-GWAS-supported *PTPRC*/CD45-mediated control of lymphocyte activation.

### Cross-ancestry heterogeneity identifies population-specific signals at 1q31

The variants with opposite AFR and EU effect directions provide additional evidence of population-specific biology at 1q31. These variants map to *LHX9, SYT2*, *ELK4*, and two intergenic positions, and five of the six show strong cross-ancestry heterogeneity. LIM homeobox 9 (*LHX9*) has been reported in lung epithelial contexts, including altered expression in alveolar epithelial type 2 cells in chronic lung disease^19^. Synaptotagmin-2 (*SYT2*) regulates calcium-dependent vesicle release, and synaptotagmin biology has been linked to regulated exocytosis in mast cells^20^, which are central effectors of IgE-mediated allergic inflammation. As an ETS-family transcription factor, *ELK4* may mark inducible gene-regulatory responses downstream of inflammatory or stress-related signaling^21^. The opposite directions may reflect population-specific effect modification. One plausible explanation is state-dependent effects, in which pathways involved in epithelial repair, secretory responses, or stress signaling are protective in acute or controlled contexts but become pathogenic under chronic inflammation^22^, potentially leading to apparent direction differences across populations with differing baseline airway states.

### Population-specific signals reveal differences in mapped functional annotations

The AFR-specific and EU-specific variant classes showed both shared and differing annotation patterns. Both classes were enriched for contractile and cytoskeletal cellular-component terms, including myofibril, sarcomere, contractile muscle fiber, supramolecular polymer, and supramolecular fiber. This shared enrichment indicates that the two ancestry-defined signal classes occupy overlapping airway-relevant biological space, rather than completely distinct pathways. These annotations are relevant to asthma because airway smooth muscle contraction, cytoskeletal remodeling, and airway-wall structural changes contribute to airway hyperresponsiveness and persistent airflow limitation^23, 24^.

The differences between the variant classes were more informative than the shared terms. The AFR-specific class showed stronger representation of contractile and calcium-regulatory annotations. The contractile annotations in this class included *CACNA1S, IGFN1, TNNT2, TNNI1, CSRP1, LMOD1, PPP1R12B, MYBPH*, and *ATP2B4*, supporting enrichment of annotations related to calcium handling, actin organization, and contractile regulation. *CACNA1S* encodes a voltage-gated calcium-channel subunit involved in excitation-contraction coupling, *LMOD1* participates in actin-filament organization in smooth muscle, and *PPP1R12B* is part of the myosin-phosphatase regulatory system involved in contractile tone^25–27^. In asthma, these features point toward the mechanical behavior of the airway wall: calcium-dependent smooth-muscle contraction, cytoskeletal remodeling, and airway narrowing^24^.

By contrast, the EU-specific class showed an added neuroaxonal annotation profile, including enrichment for axon initial segment and main axon. These terms were driven mainly by *NAV1, NFASC, CNTN2*, and *ADORA1*. *NFASC* and *CNTN2* are involved in axonal domain organization and cell-adhesion processes at specialized neuronal structures, while *NAV1* is linked to neuronal projection biology^28–30^. This suggests that EU-specific variants more strongly tag a neuroaxonal component of the same regional association landscape. Such a component is biologically plausible in asthma because parasympathetic and sensory nerves regulate bronchoconstriction, mucus secretion, cough, and airway hyperresponsiveness, and neuroimmune signaling is increasingly recognized as part of asthma pathobiology^31^. However, *ADORA1* should not be treated as uniquely EU-specific, because *ADORA1* variants were also observed in AFR. Adenosine A1 receptor (*ADORA1*) signaling has been implicated in bronchoconstriction and airway hyperresponsiveness^32^.

The cross-ancestry heterogeneous variant class further supports this interpretation. Heterogeneous variants mapped to both contractile/cytoskeletal and neuroaxonal annotations. Contractile terms included *CACNA1S, IGFN1, TNNT2, CSRP1, PPP1R12B*, and *ATP2B4*, whereas axon initial segment terms included *NAV1, NFASC*, and *CNTN2*. The supramolecular polymer and fiber terms added *NEK7*, *KIF14*, *PKP1*, and *NAV1*, placing the heterogeneous signal in a cytoskeletal and cell-structural context. *NEK7* has also been implicated in inflammasome-related biology^33^. This pattern indicates that variants with evidence of cross-ancestry effect differences highlight the same molecular patterns: airway-wall contractility, cytoskeletal organization, neuroaxonal regulation, and inflammasome. Long-term differences in infectious, allergen, and irritant exposures might have shaped the regulatory context of Th2 immune loci^34^, leading 1q31 variation to influence different airway effector pathways across populations.

### Perspective

A central contribution of this study is the demonstration that the 1q31 SDA association is highly population-specific at the variant level, despite being present in both EU and AFR ancestry groups. The small number of shared genome-wide significant variants, together with a large number of ancestry-specific associations and heterogeneous variants, reveals regional architectures that would be obscured in single-ancestry or broad asthma analyses. The results support a shared role for 1q31 in Th2-driven, glucocorticoid-dependent asthma, with population-specific differences in fine-scale genetic architecture. Variation at this locus may influence not only asthma risk, but also the biological context in which airway inflammation remains glucocorticoid-responsive. The cross-ancestry results suggest that the locus contains multiple functional components. In AFR, the signal highlights immune-cell activation together with calcium-dependent contractile and structural airway-wall biology. In EU, the enrichment pattern adds a neuroaxonal component that may connect inflammation with bronchomotor tone, cough, mucus secretion, and airway hyperresponsiveness.

Evolutionary-genetic studies have shown that immune-response regulatory variants can have ancestry-dependent effects shaped by pathogen exposure, selection, and demographic history ^35, 36^. Th2 immunity evolved in relation to parasite defense, epithelial barrier protection, and tissue repair, while also contributing to allergic disease in modern environments ^34^. More broadly, human immune variation reflects both genetic ancestry and environmental history ^37^. The present data provide a framework for future functional studies testing whether ancestry-specific regulatory architecture at 1q31 modifies the balance among immune activation, airway-wall contractility, and neurogenic airway responses in SDA.

## Acknowledgements

We thank all patients and their families who have participated in our research for the past two decades.

## Ethics approval and consent to participate

All experimental protocols were approved by the Institutional Review Board (IRB) of the Children’s Hospital of Philadelphia (CHOP) with the IRB number: IRB 16-013278. Informed consent was obtained from all subjects. If subjects are under 18, consent was also obtained from a parent and/or legal guardian with assent from the child if 7 years or older.

## Consent for publication

Not applicable.

## Competing interest

The authors declared no potential conflicts of interest with respect to the research, authorship, and/or publication of this article.

## Data availability statement

Additional information is available from the corresponding author upon request.

## Funding

The study was supported by the Institutional Development Funds from the Children’s Hospital of Philadelphia to the Center for Applied Genomics, and The Children’s Hospital of Philadelphia Endowed Chair in Genomic Research to HH.

---

**Supplementary Table 1.** The African ancestry study samples.

| Supplementary Table 1. The African ancestry study samples |  |  |  |  |  |  |  |
| --- | --- | --- | --- | --- | --- | --- | --- |
| CHIP | PHENOTYPE | Controls | MALE_1 | FEMALE_1 | Cases | MALE_2 | FEMALE_2 |
| GSA_Merge_2021 | asthma | 1109 | 648 | 461 | 139 | 64 | 75 |
| GSA_Merge_2022 | asthma | 1460 | 634 | 826 | 21 | 14 | 7 |
| GSA-24v3_2021 | asthma | 1883 | 922 | 961 | 412 | 224 | 188 |
| GSA-Merge_Dec2020_Update | asthma | 2092 | 866 | 1226 | 342 | 153 | 189 |
| GSA-Merge-subset1 | asthma | 1400 | 610 | 790 | 333 | 168 | 165 |
| GSA-Merge-subset10 | asthma | 1334 | 599 | 735 | 335 | 184 | 151 |
| GSA-Merge-subset11 | asthma | 1414 | 583 | 831 | 346 | 194 | 152 |
| GSA-Merge-subset12 | asthma | 1466 | 637 | 829 | 328 | 181 | 147 |
| GSA-Merge-subset13 | asthma | 1396 | 600 | 796 | 326 | 186 | 140 |
| GSA-Merge-subset14 | asthma | 1340 | 596 | 744 | 348 | 178 | 170 |
| GSA-Merge-subset2 | asthma | 1450 | 609 | 841 | 320 | 185 | 135 |
| GSA-Merge-subset3 | asthma | 1376 | 577 | 799 | 341 | 179 | 162 |
| GSA-Merge-subset4 | asthma | 1396 | 594 | 802 | 355 | 192 | 163 |
| GSA-Merge-subset5 | asthma | 1434 | 617 | 817 | 336 | 207 | 129 |
| GSA-Merge-subset6 | asthma | 1361 | 591 | 770 | 318 | 174 | 144 |
| GSA-Merge-subset7 | asthma | 1416 | 584 | 832 | 338 | 167 | 171 |
| GSA-Merge-subset8 | asthma | 1390 | 582 | 808 | 335 | 203 | 132 |
| GSA-Merge-subset9 | asthma | 1376 | 573 | 803 | 355 | 196 | 159 |
| HM-Merge-subset1 | asthma | 622 | 265 | 357 | 316 | 169 | 147 |
| HM-Merge-subset10 | asthma | 638 | 281 | 357 | 312 | 168 | 144 |
| HM-Merge-subset11 | asthma | 600 | 271 | 329 | 336 | 166 | 170 |
| HM-Merge-subset2 | asthma | 576 | 262 | 314 | 320 | 166 | 154 |
| HM-Merge-subset3 | asthma | 625 | 274 | 351 | 308 | 179 | 129 |
| HM-Merge-subset4 | asthma | 656 | 305 | 351 | 344 | 199 | 145 |
| HM-Merge-subset5 | asthma | 586 | 276 | 310 | 313 | 173 | 140 |
| HM-Merge-subset6 | asthma | 625 | 272 | 353 | 322 | 173 | 149 |
| HM-Merge-subset7 | asthma | 637 | 285 | 352 | 317 | 161 | 156 |
| HM-Merge-subset8 | asthma | 657 | 286 | 371 | 321 | 181 | 140 |
| HM-Merge-subset9 | asthma | 650 | 290 | 360 | 325 | 181 | 144 |
| Omni25_2021 | asthma | 600 | 258 | 342 | 273 | 145 | 128 |
| Omni25-subset1 | asthma | 496 | 334 | 162 | 172 | 100 | 72 |
| Omni25-subset2 | asthma | 485 | 290 | 195 | 189 | 130 | 59 |
| OmniExpress-OmniExpressExome-Merge-subset1 | asthma | 572 | 212 | 360 | 31 | 18 | 13 |
| OmniExpress-OmniExpressExome-Merge-subset2 | asthma | 553 | 201 | 352 | 29 | 14 | 15 |
| OmniExpress-OmniExpressExome-Merge-subset3 | asthma | 569 | 233 | 336 | 44 | 26 | 18 |
| OmniExpress-OmniExpressExome-Merge-subset4 | asthma | 576 | 223 | 353 | 36 | 22 | 14 |
| OmniExpress-OmniExpressExome-Merge-subset5 | asthma | 575 | 216 | 359 | 29 | 17 | 12 |
| GSA_Merge_2021 | NSDA | 1109 | 648 | 461 | 138 | 64 | 74 |
| GSA_Merge_2022 | NSDA | 1460 | 634 | 826 | 21 | 14 | 7 |
| GSA-24v3_2021 | NSDA | 1883 | 922 | 961 | 407 | 222 | 185 |
| GSA-Merge_Dec2020_Update | NSDA | 2092 | 866 | 1226 | 341 | 153 | 188 |
| GSA-Merge-subset1 | NSDA | 1400 | 610 | 790 | 327 | 166 | 161 |
| GSA-Merge-subset10 | NSDA | 1334 | 599 | 735 | 332 | 182 | 150 |
| GSA-Merge-subset11 | NSDA | 1414 | 583 | 831 | 343 | 192 | 151 |
| GSA-Merge-subset12 | NSDA | 1466 | 637 | 829 | 324 | 178 | 146 |
| GSA-Merge-subset13 | NSDA | 1396 | 600 | 796 | 322 | 184 | 138 |
| GSA-Merge-subset14 | NSDA | 1340 | 596 | 744 | 343 | 175 | 168 |
| GSA-Merge-subset2 | NSDA | 1450 | 609 | 841 | 317 | 183 | 134 |
| GSA-Merge-subset3 | NSDA | 1376 | 577 | 799 | 339 | 179 | 160 |
| GSA-Merge-subset4 | NSDA | 1396 | 594 | 802 | 352 | 190 | 162 |
| GSA-Merge-subset5 | NSDA | 1434 | 617 | 817 | 335 | 206 | 129 |
| GSA-Merge-subset6 | NSDA | 1361 | 591 | 770 | 314 | 174 | 140 |
| GSA-Merge-subset7 | NSDA | 1416 | 584 | 832 | 334 | 166 | 168 |
| GSA-Merge-subset8 | NSDA | 1390 | 582 | 808 | 332 | 201 | 131 |
| GSA-Merge-subset9 | NSDA | 1376 | 573 | 803 | 350 | 193 | 157 |
| HM-Merge-subset1 | NSDA | 622 | 265 | 357 | 310 | 167 | 143 |
| HM-Merge-subset10 | NSDA | 638 | 281 | 357 | 307 | 165 | 142 |
| HM-Merge-subset11 | NSDA | 600 | 271 | 329 | 336 | 166 | 170 |
| HM-Merge-subset2 | NSDA | 576 | 262 | 314 | 315 | 163 | 152 |
| HM-Merge-subset3 | NSDA | 625 | 274 | 351 | 303 | 176 | 127 |
| HM-Merge-subset4 | NSDA | 656 | 305 | 351 | 340 | 197 | 143 |
| HM-Merge-subset5 | NSDA | 586 | 276 | 310 | 307 | 170 | 137 |
| HM-Merge-subset6 | NSDA | 625 | 272 | 353 | 315 | 170 | 145 |
| HM-Merge-subset7 | NSDA | 637 | 285 | 352 | 312 | 159 | 153 |
| HM-Merge-subset8 | NSDA | 657 | 286 | 371 | 315 | 178 | 137 |
| HM-Merge-subset9 | NSDA | 650 | 290 | 360 | 322 | 180 | 142 |
| Omni25_2021 | NSDA | 600 | 258 | 342 | 270 | 144 | 126 |
| Omni25-subset1 | NSDA | 496 | 334 | 162 | 172 | 100 | 72 |
| Omni25-subset2 | NSDA | 485 | 290 | 195 | 189 | 130 | 59 |
| OmniExpress-OmniExpressExome-Merge-subset1 | NSDA | 572 | 212 | 360 | 30 | 17 | 13 |
| OmniExpress-OmniExpressExome-Merge-subset2 | NSDA | 553 | 201 | 352 | 29 | 14 | 15 |
| OmniExpress-OmniExpressExome-Merge-subset3 | NSDA | 569 | 233 | 336 | 44 | 26 | 18 |
| OmniExpress-OmniExpressExome-Merge-subset4 | NSDA | 576 | 223 | 353 | 35 | 22 | 13 |
| OmniExpress-OmniExpressExome-Merge-subset5 | NSDA | 575 | 216 | 359 | 27 | 16 | 11 |
| GSA_Merge_2021 | SDA | 1109 | 648 | 461 | 1 | 0 | 1 |
| GSA_Merge_2022 | SDA | 1460 | 634 | 826 | 0 | 0 | 0 |
| GSA-24v3_2021 | SDA | 1883 | 922 | 961 | 5 | 2 | 3 |
| GSA-Merge_Dec2020_Update | SDA | 2092 | 866 | 1226 | 1 | 0 | 1 |
| GSA-Merge-subset1 | SDA | 1400 | 610 | 790 | 6 | 2 | 4 |
| GSA-Merge-subset10 | SDA | 1334 | 599 | 735 | 3 | 2 | 1 |
| GSA-Merge-subset11 | SDA | 1414 | 583 | 831 | 3 | 2 | 1 |
| GSA-Merge-subset12 | SDA | 1466 | 637 | 829 | 4 | 3 | 1 |
| GSA-Merge-subset13 | SDA | 1396 | 600 | 796 | 4 | 2 | 2 |
| GSA-Merge-subset14 | SDA | 1340 | 596 | 744 | 5 | 3 | 2 |
| GSA-Merge-subset2 | SDA | 1450 | 609 | 841 | 3 | 2 | 1 |
| GSA-Merge-subset3 | SDA | 1376 | 577 | 799 | 2 | 0 | 2 |
| GSA-Merge-subset4 | SDA | 1396 | 594 | 802 | 3 | 2 | 1 |
| GSA-Merge-subset5 | SDA | 1434 | 617 | 817 | 1 | 1 | 0 |
| GSA-Merge-subset6 | SDA | 1361 | 591 | 770 | 4 | 0 | 4 |
| GSA-Merge-subset7 | SDA | 1416 | 584 | 832 | 4 | 1 | 3 |
| GSA-Merge-subset8 | SDA | 1390 | 582 | 808 | 3 | 2 | 1 |
| GSA-Merge-subset9 | SDA | 1376 | 573 | 803 | 5 | 3 | 2 |
| HM-Merge-subset1 | SDA | 622 | 265 | 357 | 6 | 2 | 4 |
| HM-Merge-subset10 | SDA | 638 | 281 | 357 | 5 | 3 | 2 |
| HM-Merge-subset11 | SDA | 600 | 271 | 329 | 0 | 0 | 0 |
| HM-Merge-subset2 | SDA | 576 | 262 | 314 | 5 | 3 | 2 |
| HM-Merge-subset3 | SDA | 625 | 274 | 351 | 5 | 3 | 2 |
| HM-Merge-subset4 | SDA | 656 | 305 | 351 | 4 | 2 | 2 |
| HM-Merge-subset5 | SDA | 586 | 276 | 310 | 6 | 3 | 3 |
| HM-Merge-subset6 | SDA | 625 | 272 | 353 | 7 | 3 | 4 |
| HM-Merge-subset7 | SDA | 637 | 285 | 352 | 5 | 2 | 3 |
| HM-Merge-subset8 | SDA | 657 | 286 | 371 | 6 | 3 | 3 |
| HM-Merge-subset9 | SDA | 650 | 290 | 360 | 3 | 1 | 2 |
| Omni25_2021 | SDA | 600 | 258 | 342 | 3 | 1 | 2 |
| Omni25-subset1 | SDA | 496 | 334 | 162 | 0 | 0 | 0 |
| Omni25-subset2 | SDA | 485 | 290 | 195 | 0 | 0 | 0 |
| OmniExpress-OmniExpressExome-Merge-subset1 | SDA | 572 | 212 | 360 | 1 | 1 | 0 |
| OmniExpress-OmniExpressExome-Merge-subset2 | SDA | 553 | 201 | 352 | 0 | 0 | 0 |
| **OmniExpress-OmniExpressExome-Merge-subset3** | **SDA** | **569** | **233** | **336** | **0** | **0** | **0** |
| **OmniExpress-OmniExpressExome-Merge-subset4** | **SDA** | **576** | **223** | **353** | **1** | **0** | **1** |
| **OmniExpress-OmniExpressExome-Merge-subset5** | **SDA** | **575** | **216** | **359** | **2** | **1** | **1** |

**Supplementary Table 2.** The European ancestry study samples.

| Supplementary Table 2. The European ancestry study samples |  |  |  |  |  |  |  |
| --- | --- | --- | --- | --- | --- | --- | --- |
| CHIP | PHENOTYPE | Controls | MALE_1 | FEMALE_1 | Cases | MALE_2 | FEMALE_2 |
| CytoSNP-850K-Merge | asthma | 55 | 31 | 24 | 3 | 2 | 1 |
| GSA_Merge_2021 | asthma | 4182 | 2236 | 1946 | 91 | 48 | 43 |
| GSA_Merge_2022 | asthma | 2410 | 906 | 1504 | 5 | 4 | 1 |
| GSA-24v3_2021 | asthma | 5433 | 3037 | 2396 | 142 | 82 | 60 |
| GSA-Merge_Dec2020_Update | asthma | 9275 | 4267 | 5008 | 164 | 85 | 79 |
| GSA-Merge-subset1 | asthma | 3873 | 2006 | 1867 | 162 | 107 | 55 |
| GSA-Merge-subset10 | asthma | 3819 | 2010 | 1809 | 206 | 112 | 94 |
| GSA-Merge-subset11 | asthma | 3831 | 2016 | 1815 | 208 | 116 | 92 |
| GSA-Merge-subset12 | asthma | 3809 | 2029 | 1780 | 187 | 117 | 70 |
| GSA-Merge-subset13 | asthma | 3818 | 1999 | 1819 | 213 | 117 | 96 |
| GSA-Merge-subset14 | asthma | 3846 | 2016 | 1830 | 232 | 134 | 98 |
| GSA-Merge-subset2 | asthma | 3811 | 2023 | 1788 | 213 | 119 | 94 |
| GSA-Merge-subset3 | asthma | 3853 | 2027 | 1826 | 197 | 115 | 82 |
| GSA-Merge-subset4 | asthma | 3835 | 2064 | 1771 | 210 | 115 | 95 |
| GSA-Merge-subset5 | asthma | 3863 | 2046 | 1817 | 224 | 124 | 100 |
| GSA-Merge-subset6 | asthma | 3880 | 2049 | 1831 | 220 | 132 | 88 |
| GSA-Merge-subset7 | asthma | 3845 | 2063 | 1782 | 172 | 92 | 80 |
| GSA-Merge-subset8 | asthma | 3858 | 2116 | 1742 | 190 | 109 | 81 |
| GSA-Merge-subset9 | asthma | 3832 | 2052 | 1780 | 175 | 95 | 80 |
| HM-Merge-subset1 | asthma | 1086 | 526 | 560 | 237 | 130 | 107 |
| HM-Merge-subset10 | asthma | 1161 | 575 | 586 | 218 | 123 | 95 |
| HM-Merge-subset11 | asthma | 1095 | 522 | 573 | 190 | 125 | 65 |
| HM-Merge-subset2 | asthma | 1161 | 567 | 594 | 194 | 112 | 82 |
| HM-Merge-subset3 | asthma | 1124 | 563 | 561 | 208 | 124 | 84 |
| HM-Merge-subset4 | asthma | 1093 | 527 | 566 | 221 | 127 | 94 |
| HM-Merge-subset5 | asthma | 1151 | 590 | 561 | 193 | 114 | 79 |
| HM-Merge-subset6 | asthma | 1147 | 572 | 575 | 192 | 117 | 75 |
| HM-Merge-subset7 | asthma | 1135 | 562 | 573 | 223 | 131 | 92 |
| HM-Merge-subset8 | asthma | 1162 | 550 | 612 | 182 | 108 | 74 |
| HM-Merge-subset9 | asthma | 1135 | 552 | 583 | 191 | 104 | 87 |
| Omni25_2021 | asthma | 196 | 98 | 98 | 21 | 16 | 5 |
| Omni25-subset1 | asthma | 2530 | 1542 | 988 | 79 | 59 | 20 |
| Omni25-subset2 | asthma | 2550 | 1534 | 1016 | 57 | 40 | 17 |
| OmniExpress-OmniExpressExome-Merge-subset1 | asthma | 4696 | 2342 | 2354 | 100 | 60 | 40 |
| OmniExpress-OmniExpressExome-Merge-subset2 | asthma | 4651 | 2311 | 2340 | 72 | 40 | 32 |
| OmniExpress-OmniExpressExome-Merge-subset3 | asthma | 4686 | 2420 | 2266 | 92 | 44 | 48 |
| OmniExpress-OmniExpressExome-Merge-subset4 | asthma | 4730 | 2471 | 2259 | 92 | 51 | 41 |
| OmniExpress-OmniExpressExome-Merge-subset5 | asthma | 4638 | 2333 | 2305 | 98 | 48 | 50 |
| CytoSNP-850K-Merge | NSDA | 55 | 31 | 24 | 2 | 1 | 1 |
| GSA_Merge_2021 | NSDA | 4182 | 2236 | 1946 | 89 | 46 | 43 |
| GSA_Merge_2022 | NSDA | 2410 | 906 | 1504 | 5 | 4 | 1 |
| GSA-24v3_2021 | NSDA | 5433 | 3037 | 2396 | 141 | 81 | 60 |
| GSA-Merge_Dec2020_Update | NSDA | 9275 | 4267 | 5008 | 161 | 84 | 77 |
| GSA-Merge-subset1 | NSDA | 3873 | 2006 | 1867 | 159 | 105 | 54 |
| GSA-Merge-subset10 | NSDA | 3819 | 2010 | 1809 | 198 | 110 | 88 |
| GSA-Merge-subset11 | NSDA | 3831 | 2016 | 1815 | 204 | 115 | 89 |
| GSA-Merge-subset12 | NSDA | 3809 | 2029 | 1780 | 181 | 113 | 68 |
| GSA-Merge-subset13 | NSDA | 3818 | 1999 | 1819 | 211 | 116 | 95 |
| GSA-Merge-subset14 | NSDA | 3846 | 2016 | 1830 | 227 | 130 | 97 |
| GSA-Merge-subset2 | NSDA | 3811 | 2023 | 1788 | 209 | 118 | 91 |
| GSA-Merge-subset3 | NSDA | 3853 | 2027 | 1826 | 193 | 113 | 80 |
| GSA-Merge-subset4 | NSDA | 3835 | 2064 | 1771 | 209 | 115 | 94 |
| GSA-Merge-subset5 | NSDA | 3863 | 2046 | 1817 | 218 | 120 | 98 |
| GSA-Merge-subset6 | NSDA | 3880 | 2049 | 1831 | 218 | 131 | 87 |
| GSA-Merge-subset7 | NSDA | 3845 | 2063 | 1782 | 169 | 92 | 77 |
| GSA-Merge-subset8 | NSDA | 3858 | 2116 | 1742 | 185 | 106 | 79 |
| GSA-Merge-subset9 | NSDA | 3832 | 2052 | 1780 | 170 | 93 | 77 |
| HM-Merge-subset1 | NSDA | 1086 | 526 | 560 | 228 | 124 | 104 |
| HM-Merge-subset10 | NSDA | 1161 | 575 | 586 | 205 | 118 | 87 |
| HM-Merge-subset11 | NSDA | 1095 | 522 | 573 | 178 | 118 | 60 |
| HM-Merge-subset2 | NSDA | 1161 | 567 | 594 | 186 | 108 | 78 |
| HM-Merge-subset3 | NSDA | 1124 | 563 | 561 | 202 | 121 | 81 |
| HM-Merge-subset4 | NSDA | 1093 | 527 | 566 | 210 | 120 | 90 |
| HM-Merge-subset5 | NSDA | 1151 | 590 | 561 | 184 | 110 | 74 |
| HM-Merge-subset6 | NSDA | 1147 | 572 | 575 | 184 | 111 | 73 |
| HM-Merge-subset7 | NSDA | 1135 | 562 | 573 | 213 | 128 | 85 |
| HM-Merge-subset8 | NSDA | 1162 | 550 | 612 | 173 | 103 | 70 |
| HM-Merge-subset9 | NSDA | 1135 | 552 | 583 | 183 | 100 | 83 |
| Omni25_2021 | NSDA | 196 | 98 | 98 | 21 | 16 | 5 |
| Omni25-subset1 | NSDA | 2530 | 1542 | 988 | 77 | 57 | 20 |
| Omni25-subset2 | NSDA | 2550 | 1534 | 1016 | 56 | 39 | 17 |
| OmniExpress-OmniExpressExome-Merge-subset1 | NSDA | 4696 | 2342 | 2354 | 97 | 58 | 39 |
| OmniExpress-OmniExpressExome-Merge-subset2 | NSDA | 4651 | 2311 | 2340 | 69 | 39 | 30 |
| OmniExpress-OmniExpressExome-Merge-subset3 | NSDA | 4686 | 2420 | 2266 | 92 | 44 | 48 |
| OmniExpress-OmniExpressExome-Merge-subset4 | NSDA | 4730 | 2471 | 2259 | 88 | 49 | 39 |
| OmniExpress-OmniExpressExome-Merge-subset5 | NSDA | 4638 | 2333 | 2305 | 93 | 46 | 47 |
| CytoSNP-850K-Merge | SDA | 55 | 31 | 24 | 1 | 1 | 0 |
| GSA_Merge_2021 | SDA | 4182 | 2236 | 1946 | 2 | 2 | 0 |
| GSA_Merge_2022 | SDA | 2410 | 906 | 1504 | 0 | 0 | 0 |
| GSA-24v3_2021 | SDA | 5433 | 3037 | 2396 | 1 | 1 | 0 |
| GSA-Merge_Dec2020_Update | SDA | 9275 | 4267 | 5008 | 3 | 1 | 2 |
| GSA-Merge-subset1 | SDA | 3873 | 2006 | 1867 | 3 | 2 | 1 |
| GSA-Merge-subset10 | SDA | 3819 | 2010 | 1809 | 8 | 2 | 6 |
| GSA-Merge-subset11 | SDA | 3831 | 2016 | 1815 | 4 | 1 | 3 |
| GSA-Merge-subset12 | SDA | 3809 | 2029 | 1780 | 6 | 4 | 2 |
| GSA-Merge-subset13 | SDA | 3818 | 1999 | 1819 | 2 | 1 | 1 |
| GSA-Merge-subset14 | SDA | 3846 | 2016 | 1830 | 5 | 4 | 1 |
| GSA-Merge-subset2 | SDA | 3811 | 2023 | 1788 | 4 | 1 | 3 |
| GSA-Merge-subset3 | SDA | 3853 | 2027 | 1826 | 4 | 2 | 2 |
| GSA-Merge-subset4 | SDA | 3835 | 2064 | 1771 | 1 | 0 | 1 |
| GSA-Merge-subset5 | SDA | 3863 | 2046 | 1817 | 6 | 4 | 2 |
| GSA-Merge-subset6 | SDA | 3880 | 2049 | 1831 | 2 | 1 | 1 |
| GSA-Merge-subset7 | SDA | 3845 | 2063 | 1782 | 3 | 0 | 3 |
| GSA-Merge-subset8 | SDA | 3858 | 2116 | 1742 | 5 | 3 | 2 |
| GSA-Merge-subset9 | SDA | 3832 | 2052 | 1780 | 5 | 2 | 3 |
| HM-Merge-subset1 | SDA | 1086 | 526 | 560 | 9 | 6 | 3 |
| HM-Merge-subset10 | SDA | 1161 | 575 | 586 | 13 | 5 | 8 |
| HM-Merge-subset11 | SDA | 1095 | 522 | 573 | 12 | 7 | 5 |
| HM-Merge-subset2 | SDA | 1161 | 567 | 594 | 8 | 4 | 4 |
| HM-Merge-subset3 | SDA | 1124 | 563 | 561 | 6 | 3 | 3 |
| HM-Merge-subset4 | SDA | 1093 | 527 | 566 | 11 | 7 | 4 |
| HM-Merge-subset5 | SDA | 1151 | 590 | 561 | 9 | 4 | 5 |
| HM-Merge-subset6 | SDA | 1147 | 572 | 575 | 8 | 6 | 2 |
| HM-Merge-subset7 | SDA | 1135 | 562 | 573 | 10 | 3 | 7 |
| HM-Merge-subset8 | SDA | 1162 | 550 | 612 | 9 | 5 | 4 |
| HM-Merge-subset9 | SDA | 1135 | 552 | 583 | 8 | 4 | 4 |
| Omni25_2021 | SDA | 196 | 98 | 98 | 0 | 0 | 0 |
| Omni25-subset1 | SDA | 2530 | 1542 | 988 | 2 | 2 | 0 |
| Omni25-subset2 | SDA | 2550 | 1534 | 1016 | 1 | 1 | 0 |
| OmniExpress-OmniExpressExome-Merge-subset1 | SDA | 4696 | 2342 | 2354 | 3 | 2 | 1 |
| OmniExpress-OmniExpressExome-Merge-subset2 | SDA | 4651 | 2311 | 2340 | 3 | 1 | 2 |
| OmniExpress-OmniExpressExome-Merge-subset3 | SDA | 4686 | 2420 | 2266 | 0 | 0 | 0 |
| OmniExpress-OmniExpressExome-Merge-subset4 | SDA | 4730 | 2471 | 2259 | 4 | 2 | 2 |
| OmniExpress-OmniExpressExome-Merge-subset5 | SDA | 4638 | 2333 | 2305 | 5 | 2 | 3 |

| Column | Meaning |
| --- | --- |
| CHIP | Chip / input dataset name |
| PHENOTYPE | Phenotype being summarized |
| N_COVAR | Number of samples in the chip-specific covariate file |
| N_PHENO_NONMISSING | Number of matched samples with non-missing phenotype |
| N_MALE | Number of non-missing phenotype samples with SEX=1 |
| N_FEMALE | Number of non-missing phenotype samples with SEX=2 |
| N_UNKNOWN_SEX | Number of non-missing phenotype samples with sex not coded as 1 or 2 |
| PHENO_VALUE_COUNTS | Counts of each observed non-missing phenotype value |
| MALE_1 | Number of males with phenotype value 1 |
| FEMALE_1 | Number of females with phenotype value 1 |
| UNKNOWN_1 | Number of unknown-sex samples with phenotype value 1 |
| MALE_2 | Number of males with phenotype value 2 |
| FEMALE_2 | Number of females with phenotype value 2 |
| UNKNOWN_2 | Number of unknown-sex samples with phenotype value 2 |

## Notes

### Competing Interest Statement

The authors have declared no competing interest.

### Author Declarations

All experimental protocols were approved by the Institutional Review Board (IRB) of the Children's Hospital of Philadelphia (CHOP) with the IRB number: IRB 16-013278.

